# The Detection of Xylazine in Tijuana, Mexico: Triangulating Drug Checking and Clinical Urine Testing Data

**DOI:** 10.1101/2024.08.19.24312273

**Authors:** Joseph R. Friedman, Alejando González Montoya, Carmina Ruiz, Mariana Alejandra González Tejeda, Luis A. Segovia, Morgan E. Godvin, Edward Sisco, Elise M. Pyfrom, Meghan G. Appley, Chelsea L. Shover, Lilia Pacheco Bufanda

## Abstract

**Introduction:** Xylazine is a veterinary anesthetic increasingly present alongside illicit fentanyl in the US and Canada, presenting novel health risks. Although xylazine remains less common in the Western US, Mexican border cities serve as key trafficking hubs and may have higher prevalence of novel substances, but surveillance has been limited.

**Methods:** We examined deidentified records from the Prevencasa harm reduction clinic in Tijuana, describing urine and paraphernalia testing from patients reporting using illicit opioids within 24 hr. Xylazine (two types), fentanyl, opiate, methamphetamine, amphetamine, benzodiazepine, and nitazene test strips were used to test urine and paraphernalia samples. Paraphernalia samples were also analyzed with mass spectrometry.

**Results:** The study consisted of 23 participants that provided both urine and paraphernalia samples. Of the participants studied, 100 %, 91.3 %, and 69.6 % reported using China White/fentanyl, methamphetamine, and tar heroin, respectively. The mean age was 41.7 years, 95.7 % were male, 65.2 % were unhoused, and 30.4 % had skin wounds at the time of sample collection.

Xylazine positivity in urine, for the two types used, was 82.6 % and 65.2 %. For paraphernalia testing, the xylazine positivity was 65.2 % and 47.8 %. Confirmatory testing of paraphernalia samples by mass spectrometry indicated a 52.2 % xylazine positivity. This testing also revealed positivity rates for fentanyl (73.9 %), fluorofentanyl (30.4 %), tramadol (30.4 %), and lidocaine (30.4 %).

The mass spectrometry results suggest lidocaine triggered *n* = 3 and *n* = 0 false positives among the xylazine test strip types. A total of *n* = 0 and *n* = 1 false negatives were also observed.

**Discussion:** Xylazine is present on the U.S.-Mexico border, requiring public health intervention. High lidocaine positivity complicates the clinical detection of xylazine via testing strips. Xylazine was found to be more prevalent in urine than in paraphernalia samples. Confirmatory urine studies are needed to better understand possible complications of using test strips for toxicological testing.

## Introduction

Xylazine is an alpha-2 adrenergic agonist, not approved for human use, that is commonly utilized in veterinary medicine for sedating animals (Hoffmann et al., 2001). Xylazine emerged as an additive to the illicit heroin supply in Puerto Rico in the early 2000s (Reyes et al., 2012), and subsequently rose to prominence as an agent used to augment illicit fentanyl in Philadelphia in the late 2010s (Johnson et al., 2021). It has become increasingly prevalent alongside illicit fentanyl across the U.S. and Canada, and has been associated with novel health risks for people who use drugs (PWUD) (Cano et al., 2024). As a potent, short-acting sedative, xylazine use has been associated with increased risk of unwanted sedation and associated vulnerability to theft, physical and sexual assault, and shifting overdose risk (e.g., sedation not responsive to naloxone) (Friedman et al., 2022c). Xylazine has also long been associated with increased risk of skin and soft tissue wounds that are distinct from typical injection-related abscesses (Dowton et al., 2023).

Xylazine prevalence in the U.S. remains higher in the Northeast, Southeast, and Midwest, and lower in the West, including California and other states on the U.S. Southern border (Cano et al., 2024). Though drug trends along the Mexican border may mimic the U.S. Southern border, cities in this area serve as key trafficking hubs and may have higher prevalence of novel substances, such as xylazine (Friedman et al., 2022a). In the context of limited epidemiological surveillance, little academic literature has focused on xylazine’s potential presence in Mexico.

The clinical and epidemiological detection of xylazine in Mexico has been complicated by limited routine testing. Xylazine test strips offer an inexpensive point-of-care option, providing a binary present/absent result, and have proven broadly reliable for analysis of actual drug product or paraphernalia (Sisco et al., 2024). Studies have also demonstrated that xylazine test strips can be used on urine samples (Hauschild et al., 2023), however, no work has looked at comparing drug checking and clinical testing results from the same person.

Here we use records from a harm reduction and free medical clinic in Tijuana, Mexico to 1) investigate the prevalence of xylazine on the U.S.-Mexico border in 2024, 2) understand the utility of xylazine test strips for urine and paraphernalia testing, and 3) determine the correlation between paired urine and paraphernalia test strip for individuals. We also conducted confirmatory mass spectrometry analysis on the paraphernalia samples to aid in understanding the limitations of the xylazine test strips. Finally, we employed a series of other drug test strips, in additional to xylazine test strips, on both urine and paraphernalia samples to determine the correlation between sample types and between test strip and confirmatory mass spectrometry results.

## Methods

We analyzed deidentified records from Prevencasa A.C., a syringe exchange program in Tijuana, Mexico which offers no-cost drug checking and clinical services for a highly vulnerable population of people who use drugs, many of whom have been deported from the United States. The clinic is located in the Zona Norte neighborhood of Tijuana, which concentrates a high volume of drug sales points, drug and sex tourism, as well as unhoused and refugee populations (Friedman et al., 2022b).

Deidentified records were provided by the clinical team from patients seen in 2024, who reported opioid use within the past 24 hr., and who participated in both drug checking services and clinical urine screening on the same day. Urine testing included immunoassay test strips for detection of several common and novel synthetic illicit substances including xylazine, fentanyl, opiate, methamphetamine, amphetamine, and benzodiazepine. Two types of xylazine test strips were used, Wisebatch (Costa Mesa, CA, USA) and SAFElife (Dayton, MT, USA), that have manufacturer reported detection thresholds of 1000 ng/mL and 500 ng/mL, respectively. Urine tests were provided in a point-of-care manner, with results reported to patients during the clinical visit.

Drug checking was performed by sampling paraphernalia (i.e., wrappers or bags) that came into direct contact with drug materials. First, a sample for confirmatory mass spectrometry analysis was obtained by swabbing both sides of a wrapper or baggie with a cotton-tipped applicator swab which was then placed in an envelope and sent to the laboratory. Subsequently, each wrapper or bag was submerged in 5 mL of sterile water and removed. The same panel of test strips as the urine analysis was used to analyze the aqueous solution. Nitazene test strips were also employed here. Each test strip was submerged to the indicator line and read after 5 min, following the manufacturer’s recommendations. In the very rare case of an invalid or indeterminate result, a second test strip was used.

Test strip results from the paraphernalia samples were corroborated with confirmatory results from direct analysis in real time mass spectrometry (DART-MS) completed at the National Institute of Standards and Technology (NIST). No identifiable information was provided to NIST scientists. Detailed protocols are provided elsewhere (Appley et al., 2023), but, in brief, residue samples on the cotton-tipped applicator swabs were extracted using acetonitrile and the resulting solutions were analyzed on a JEOL 4G AccuTOF (Peabody, MA, USA) coupled with a Bruker DART-SVP ion source (Billerica, MA, USA). Two in-source collision induced dissociation mass spectra for each sample were obtained and were searched against an in-house library containing over 1,300 drugs, cutting agents, and adulterants for identification using a minimum 3 % relative abundance threshold and a ±5 mDa mass tolerance threshold. For a small number of samples where the test strip result indicated xylazine positivity that was not detected by DART-MS, an additional analysis of the acetonitrile solution was completed using liquid chromatography tandem mass spectrometry (LC-MS/MS) (Thermo UltiMate 3000 (Waltham, MA, USA) coupled with a Sciex QTrap 4000 (Framingham, MA, USA)). The system was operated in multi-reaction mode, using analytical methods discussed in detail elsewhere (Sisco et al., 2023), to determine if xylazine was present in the solution at a level that was below the limit of detection for the DART-MS method.

## Results

The samples consisted of *n* = 23 patients with both urine and paraphernalia tests completed (Table 1). The mean age was 41.7, *n* = 22 (95.7 %) were male, *n* = 15 (65.2 %) were unhoused, *n* = 11 (47.8 %) had previously been deported, and *n* = 7 (30.4 %) had skin wounds at the time of their clinical encounter. Of the participants, *n* = 23 (100 %), *n* = 21 (91.3 %), and *n* = 16 (69.6 %) reported using China White/fentanyl, methamphetamine, and black tar heroin, respectively (Table 1).

**Table 1.**
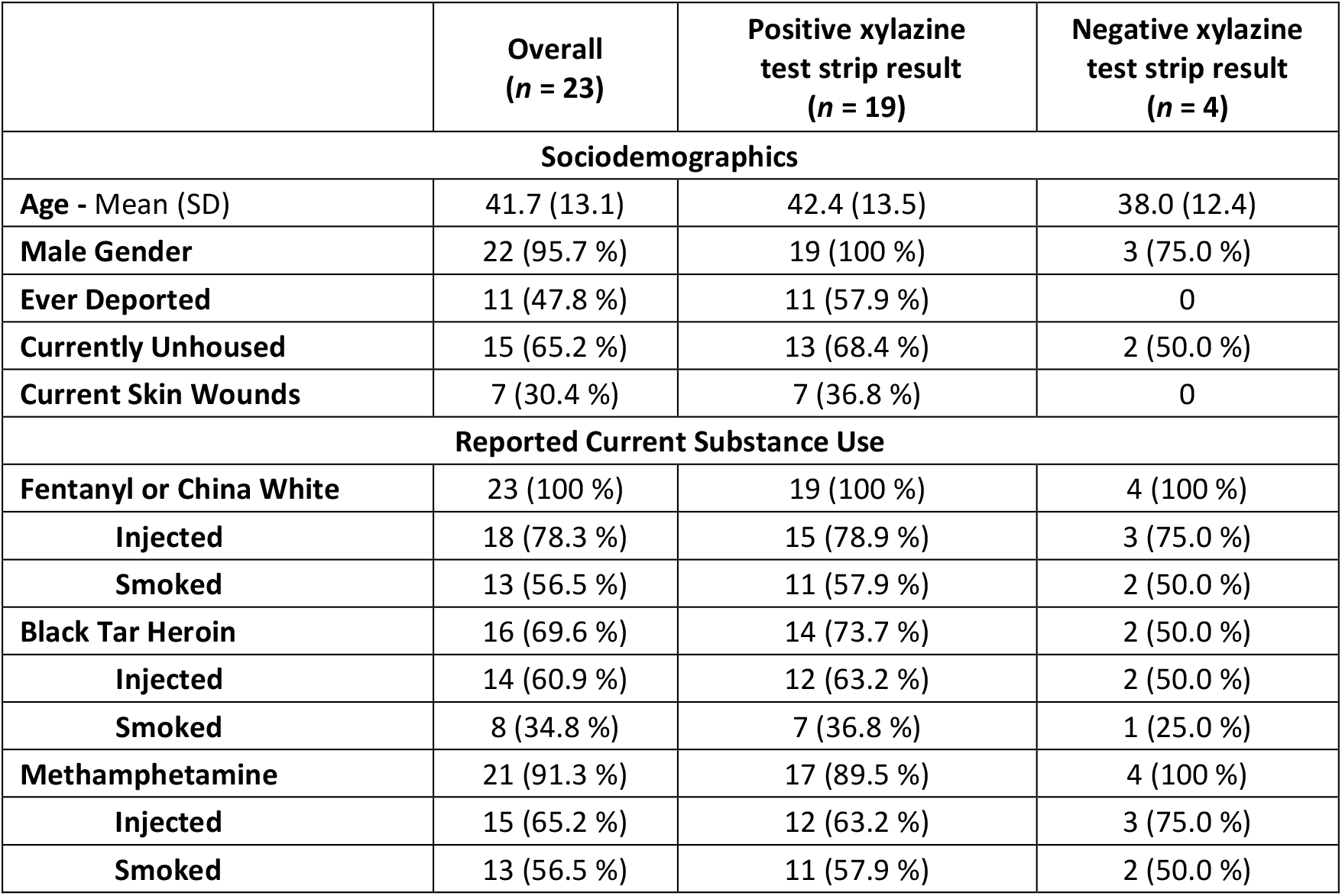
Participant demographics and reported substance use statistics. Summary statistics as well as statistics broken out by the urine xylazine test strip result (using Wisebatch test strips) are provided.

| | Overall<br>($n = 23$) | Positive xylazine<br>test strip result<br>($n = 19$) | Negative xylazine<br>test strip result<br>($n = 4$) |
| --- | --- | --- | --- |
| **Sociodemographics** |  |  |  |
| Age - Mean (SD) | 41.7 (13.1) | 42.4 (13.5) | 38.0 (12.4) |
| Male Gender | 22 (95.7 %) | 19 (100 %) | 3 (75.0 %) |
| Ever Deported | 11 (47.8 %) | 11 (57.9 %) | 0 |
| Currently Unhoused | 15 (65.2 %) | 13 (68.4 %) | 2 (50.0 %) |
| Current Skin Wounds | 7 (30.4 %) | 7 (36.8 %) | 0 |
| **Reported Current Substance Use** |  |  |  |
| Fentanyl or China White | 23 (100 %) | 19 (100 %) | 4 (100 %) |
| Injected | 18 (78.3 %) | 15 (78.9 %) | 3 (75.0 %) |
| Smoked | 13 (56.5 %) | 11 (57.9 %) | 2 (50.0 %) |
| Black Tar Heroin | 16 (69.6 %) | 14 (73.7 %) | 2 (50.0 %) |
| Injected | 14 (60.9 %) | 12 (63.2 %) | 2 (50.0 %) |
| Smoked | 8 (34.8 %) | 7 (36.8 %) | 1 (25.0 %) |
| Methamphetamine | 21 (91.3 %) | 17 (89.5 %) | 4 (100 %) |
| Injected | 15 (65.2 %) | 12 (63.2 %) | 3 (75.0 %) |
| Smoked | 13 (56.5 %) | 11 (57.9 %) | 2 (50.0 %) |

Urine xylazine positivity was *n* = 19 (82.6 %) (Table 1) and *n* = 15 (65.2 %) using Wisebatch and SAFElife test strips, respectively. Urine test strip positivity was *n* = 23 (100 %) for fentanyl, *n* = 22 (95.7 %) for methamphetamine, *n* = 10 (43.5 %) for opiates, and *n* = 2 (8.7 %) for benzodiazepines (Table 2).

**Table 2.**
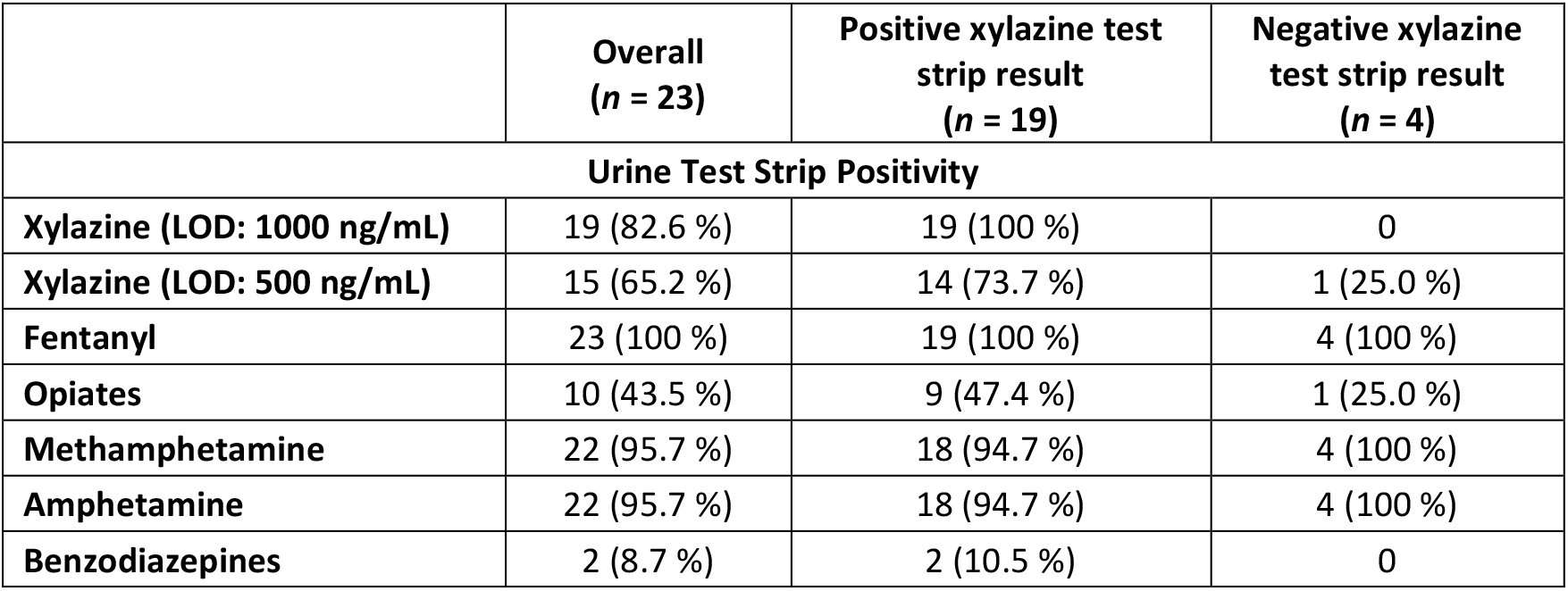
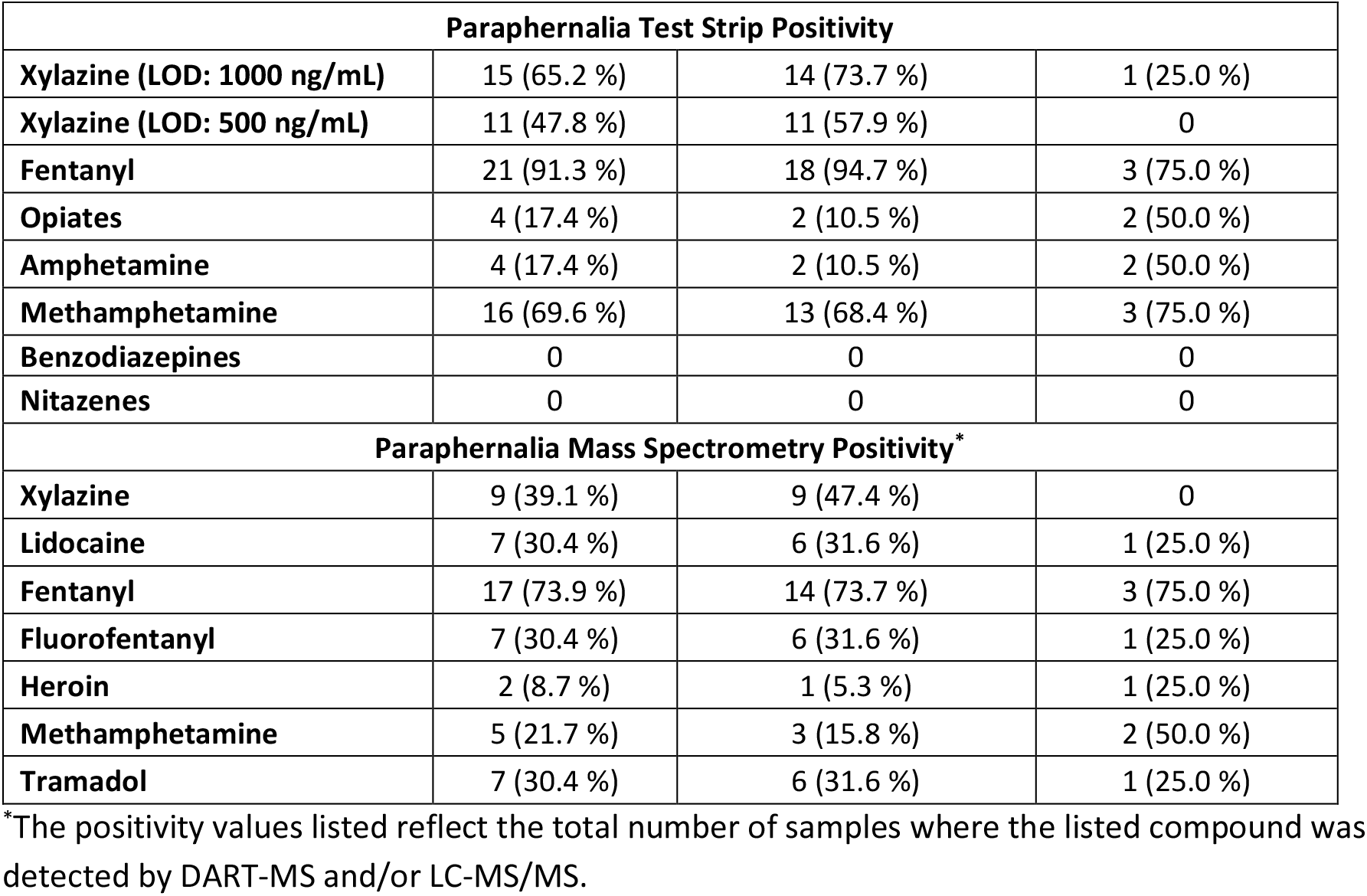
Summary of positivity results for urine and paraphernalia tests conducted. Overall information as well as information broken out by urine xylazine test strip result (using Wisebatch test strips) is provided.

| | Overall<br>($n = 23$) | Positive xylazine test<br>strip result<br>($n = 19$) | Negative xylazine<br>test strip result<br>($n = 4$) |
| --- | --- | --- | --- |
| **Urine Test Strip Positivity** |  |  |  |
| Xylazine (LOD: 1000 ng/mL) | 19 (82.6 %) | 19 (100 %) | 0 |
| Xylazine (LOD: 500 ng/mL) | 15 (65.2 %) | 14 (73.7 %) | 1 (25.0 %) |
| Fentanyl | 23 (100 %) | 19 (100 %) | 4 (100 %) |
| Opiates | 10 (43.5 %) | 9 (47.4 %) | 1 (25.0 %) |
| Methamphetamine | 22 (95.7 %) | 18 (94.7 %) | 4 (100 %) |
| Amphetamine | 22 (95.7 %) | 18 (94.7 %) | 4 (100 %) |
| Benzodiazepines | 2 (8.7 %) | 2 (10.5 %) | 0 |

| Paraphernalia Test Strip Positivity |  |  |  |
| --- | --- | --- | --- |
| Xylazine (LOD: 1000 ng/mL) | 15 (65.2 %) | 14 (73.7 %) | 1 (25.0 %) |
| Xylazine (LOD: 500 ng/mL) | 11 (47.8 %) | 11 (57.9 %) | 0 |
| Fentanyl | 21 (91.3 %) | 18 (94.7 %) | 3 (75.0 %) |
| Opiates | 4 (17.4 %) | 2 (10.5 %) | 2 (50.0 %) |
| Amphetamine | 4 (17.4 %) | 2 (10.5 %) | 2 (50.0 %) |
| Methamphetamine | 16 (69.6 %) | 13 (68.4 %) | 3 (75.0 %) |
| Benzodiazepines | 0 | 0 | 0 |
| Nitazenes | 0 | 0 | 0 |
| Paraphernalia Mass Spectrometry Positivity* |  |  |  |
| Xylazine | 9 (39.1 %) | 9 (47.4 %) | 0 |
| Lidocaine | 7 (30.4 %) | 6 (31.6 %) | 1 (25.0 %) |
| Fentanyl | 17 (73.9 %) | 14 (73.7 %) | 3 (75.0 %) |
| Fluorofentanyl | 7 (30.4 %) | 6 (31.6 %) | 1 (25.0 %) |
| Heroin | 2 (8.7 %) | 1 (5.3 %) | 1 (25.0 %) |
| Methamphetamine | 5 (21.7 %) | 3 (15.8 %) | 2 (50.0 %) |
| Tramadol | 7 (30.4 %) | 6 (31.6 %) | 1 (25.0 %) |

\*The positivity values listed reflect the total number of samples where the listed compound was detected by DART-MS and/or LC-MS/MS.

The paraphernalia samples were reported by the participants to have contained China White/fentanyl alone for *n* = 8 samples (34.8 %), China White/fentanyl and methamphetamine for *n* = 10 samples (43.5 %), black tar heroin alone for *n* = 4 (17.4 %) samples, and methamphetamine alone for *n* = 1 (4.3 %) sample.

Xylazine test strip positivity for paraphernalia samples was lower than urine samples, at *n* = 15 (65.2 %) and *n* = 11 (47.8 %) for Wisebatch and SAFElife strips, respectively. Paraphernalia test strip positivity was *n* = 21 (91.3 %) for fentanyl, *n* = 16 (69.6 %) for methamphetamine, *n* = 4 (17.4 %) for opiates, and 0 % for nitazenes and benzodiazepines (Table 2). The correlation and concordance between paraphernalia and urine xylazine test strip results are shown in Supplemental Figure 1 (correlation) and Supplemental Table 1 (concordance).

Confirmatory mass spectrometry analysis indicated a *n* = 12 (52.2 %) xylazine positivity in paraphernalia samples, as well as *n* = 17 (73.9 %) for fentanyl, *n* = 7 (30.4 %) for fluorofentanyl, *n* = 7 (30.4 %) for tramadol, and *n* = 7 (30.4 %) for lidocaine. A summary of all individual results is provided in Figure 1. Comparing xylazine positivity for paraphernalia test strips (Wisebatch) and mass spectrometry indicates 12 true positives, eight true negatives, three false positives, and zero false negatives (Supplemental Table 1). For each of the three false positives, lidocaine (known to cross-react with xylazine strips) was detected by mass spectrometry. For SAFElife strips, 11 true positives, 11 true negatives, zero false positives, and one false negative was observed. Correlation coefficients with mass spectrometry, were r = 0.76 and r = 0.92 for Wisebatch and SAFElife xylazine test strips, respectively.

**Figure 1.**
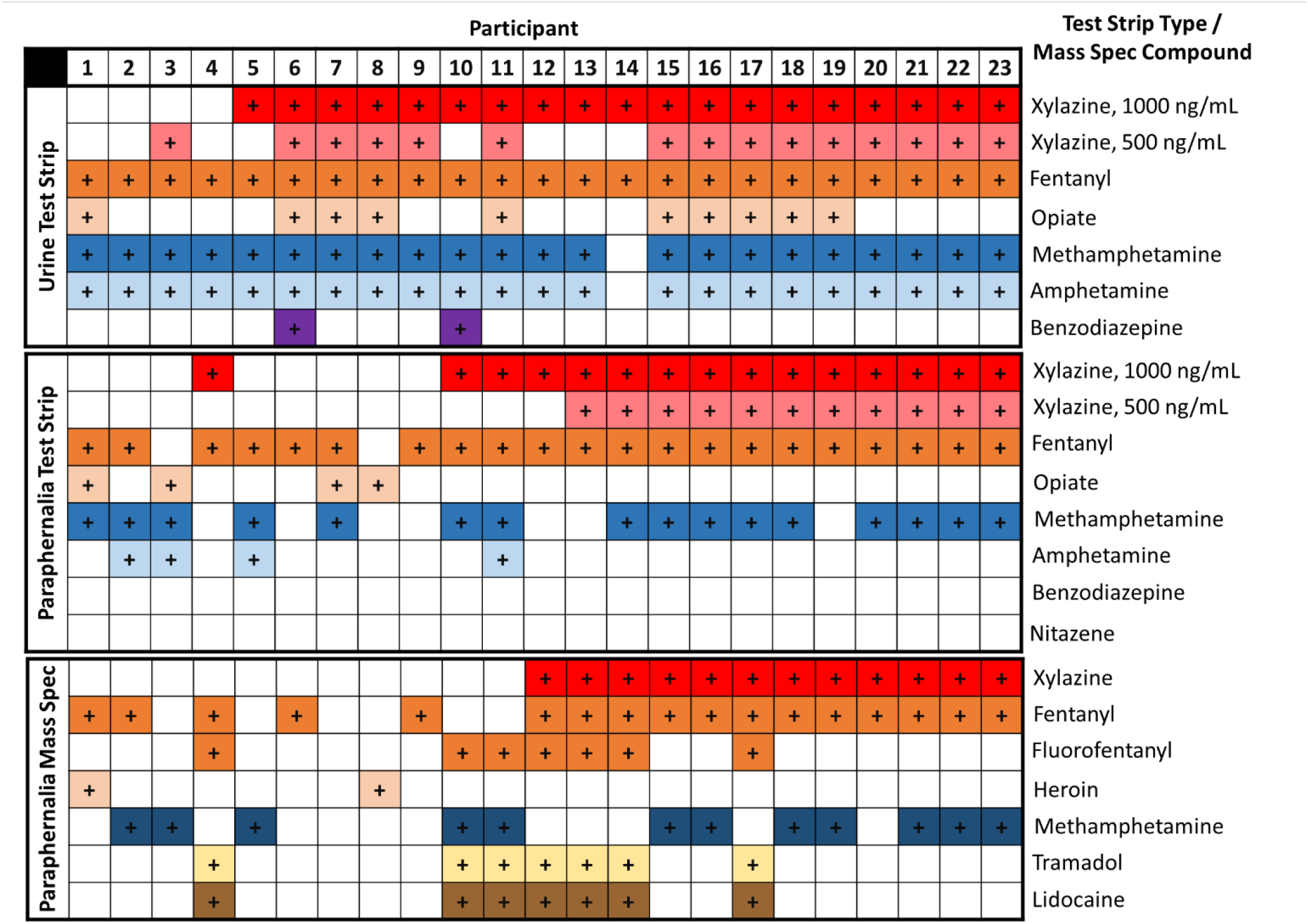
Participant-level data. Each column describes, for a single participant, from top to bottom, urine test strip results, paraphernalia test strip results, and paraphernalia mass spectrometry results. A colored cell with a “+” indicates a positive result.

Concordance and correlation between urine test strip and paraphernalia mass spectrometry results from the same individuals were lower, reflecting higher xylazine positivity within urine samples. There were no instances in which Wisebatch test strips analyzing urine produced a negative result when mass spectrometry paraphernalia results indicated the presence of xylazine. However, for *n* = 7 individuals, urine tests were positive when paraphernalia tests were negative. The correlation coefficient between Wisebatch test strip urine results and mass spectrometry paraphernalia testing for xylazine was r = 0.48. SAFElife test strip results for urine samples were negative on three occasions where paraphernalia mass spectrometry results were positive for xylazine. This relationship showed a correlation coefficient of r = 0.21.

## Discussion

In this small exploratory study, we provide the first epidemiological evidence in the academic literature, to our knowledge, of xylazine’s presence in the illicit drug supply on the U.S.-Mexico border. Compared to other studies describing xylazine in Western North America (Cano et al., 2024), we noted a high rate of xylazine positivity, with over 80 % of urine samples testing positive on test strips, and over 50 % of paraphernalia samples testing positive via mass spectrometry. This study also represents a methodological contribution in the linkage of clinical urine testing and direct substance analysis—an approach that may have broad utility for triangulation of health risks in harm reduction centers offering both clinical and drug checking programs.

We found that xylazine is present on the US-Mexico border, which represents novel health risks for people who use drugs. The increased risk from xylazine for overdose, soft tissue infection, unwanted sedation, and other associated health harms for people who use drugs in Mexico deserves careful attention for public health priority setting in Mexico. Given Tijuana’s strategic location on illicit drug supply lines to the US, these findings may also suggest that xylazine prevalence may soon rise in Southern California and other proximate locations to the U.S.-Mexico border.

Although this study was limited by a small sample size, one strength is the large number of clinical and drug checking indicators collected in the same individuals, allowing for assessing within-participant concordance between various methods of assessing xylazine positivity. We found a higher rate of xylazine positivity in urine samples compared to paraphernalia samples. This is perhaps intuitive, as most consumers in Zona Norte purchase samples from various sales points, therefore their urine may reflect a wider temporal window compared to a single drug sample. Nevertheless, little is known about xylazine’s active detection window in urine, and further studies are needed to better understand the utility of point-of-care xylazine urine testing over time. Another possibility that deserves consideration is that the 5 mL of water used to prepare drug wrappers may have resulted in false negative results from over-dilution. This quantity of sterile water was deemed necessary to fully wet the samples, which at times, were large wrappers difficult to fully cover with a lower volume of liquid. Nevertheless, smaller quantities of water may have resulted in a higher xylazine positivity rate in substance samples, and various dilutions could be explored in further studies.

This work also highlights potential challenges with implementation of test strips for paraphernalia and urine testing. The observed performance of the test strips was not identical, likely due to different antibodies being used, as evidenced by lidocaine cross-reactivity only occurring with one of the test strips. Also, the test strip with a lower reported sensitivity was found to have a lower positivity rate, counter to the expected result. Repeat studies with larger sample sizes are required to draw more definitive conclusions, but these results highlight the often confusing and rapidly moving landscape of point-of-care drug checking technologies and the need for increased standardization in this field.

Beyond xylazine test strips, confirmatory mass spectrometry analysis of the paraphernalia samples revealed other interesting aspects of the illicit opioid supply in Tijuana. We detected a high prevalence of fluorofentanyl, a trend noted in other jurisdictions across the U.S. (Bitting, 2022), as well as fentanyl, and lidocaine (both commonly available pharmaceutical agents in Tijuana). Tramadol is a weak opioid that is available over the counter in Mexico and may therefore be attractive as a cheap and readily available bulking agent. The high prevalence of lidocaine in the illicit fentanyl supply poses challenges for reliable clinical detection of xylazine, as it is known to trigger false positives in some generations of xylazine test strips. Although newer generations of test strips may reduce this problem, clinicians should be aware of the possibility of false positive results in areas with high lidocaine prevalence. Nevertheless, we are not aware of any other studies demonstrating a high prevalence of lidocaine use in illicit fentanyl samples, and more research in Tijuana is needed to understand the motivations for this practice.

In sum, we provide early evidence that xylazine is present on the U.S.-Mexico border, which requires public health intervention. We also highlight some of the strengths and challenges of triangulating clinical and drug checking data, and the complexities and uncertainties of point-of-case xylazine detection technologies. Although this study highlights the feasibility of using immunoassay test strips during routine clinical encounters for the detection of xylazine, confirmatory studies are needed to better describe the limitations of this approach. In particular, the high lidocaine positivity in illicit fentanyl samples in Tijuana was an unexpected complicating factor that deserves further consideration.

## Data Availability

All data produced in the present work are contained in the manuscript (in Figure 1, which shows participant-level results).

## Disclaimer

Certain commercial products are identified in order to adequately specify the procedure; this does not imply endorsement or recommendation by NIST, nor does it imply that such products are necessarily the best available for the purpose.

All test strips were purchased by the researchers, and the manufacturers played no role in the study design or interpretation. Study protocols were approved by the Institutional Review Board at Prevencasa A.C.

## Author Contributions

JRF and LBP lead the study. JRF, LBP, AGM, CLS, MG, and ES conceptualized the methods. AGM, CR, MAGT, and LS contributed to data collection and management. JRF conducted the analysis and wrote the first draft. All authors contributed to the design and interpretation of results and revision of the article.

## Supplemental Information

**Supplemental Figure 1.**
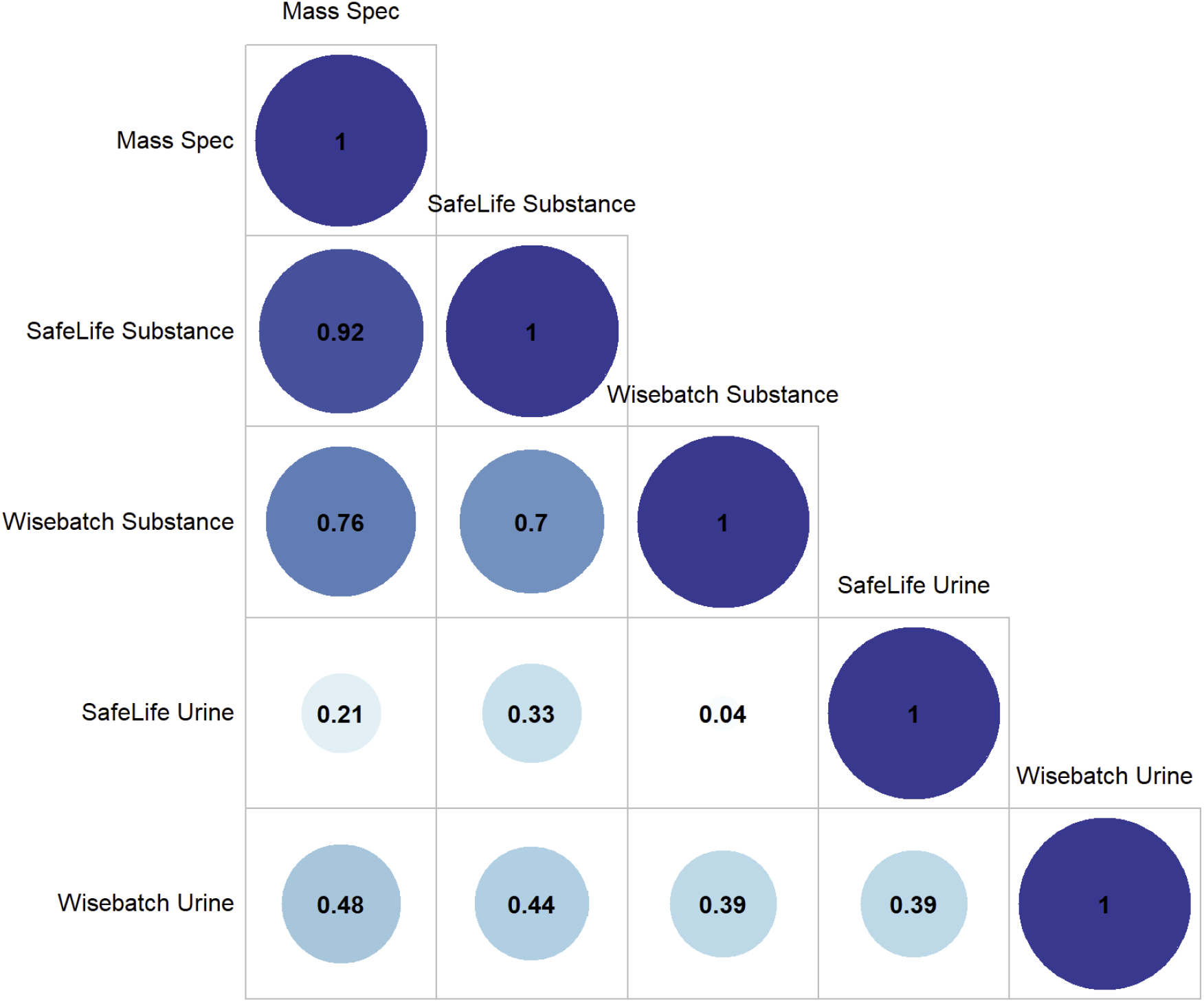
Correlation between xylazine detection among each pair of results obtain from testing the substance or urine.

**Supplemental Table 1.**
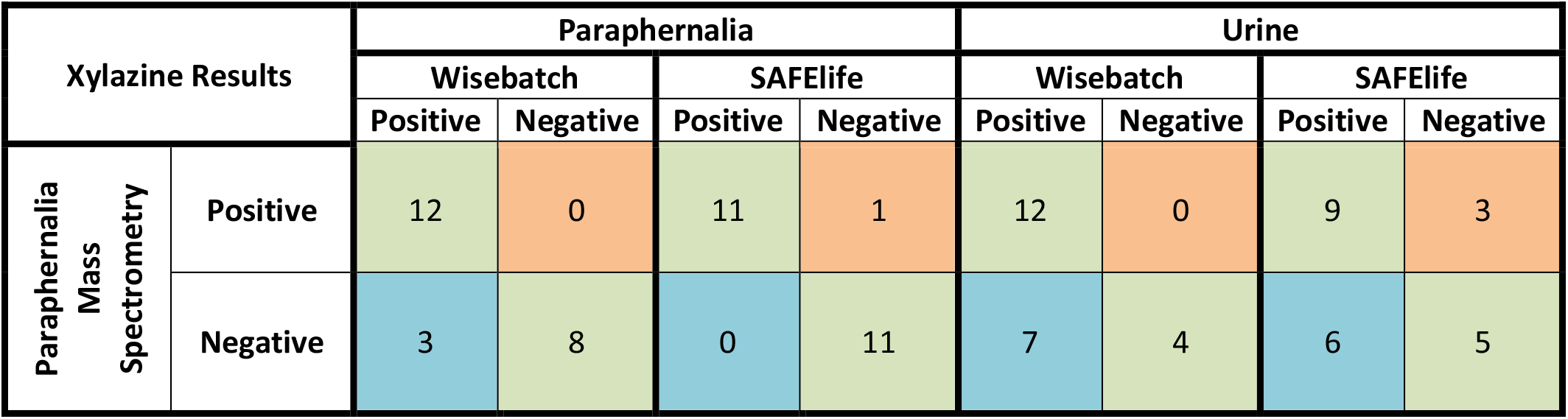
Concordance between paraphernalia mass spectrometry results and test strip results from the paraphernalia and urine samples. Green cells indicate concordant results, blue cells indicate false positive results, and orange cells indicate false negative results.

